# Symptoms and signs of lung cancer prior to diagnosis: Comparative study using electronic health records

**DOI:** 10.1101/2022.06.01.22275657

**Authors:** Maria G. Prado, Larry G. Kessler, Margaret A. Au, Hannah A. Burkhardt, Monica Zigman Suchsland, Lesleigh Kowalski, Kari A. Stephens, Meliha Yetisgen, Fiona M. Walter, Richard D. Neal, Kevin Lybarger, Caroline A. Thompson, Morhaf Al Achkar, Elizabeth A. Sarma, Grace Turner, Farhood Farjah, Matthew Thompson

## Abstract

**Background:** Lung cancer is the most common cause of cancer-related death in the United States (US), with most patients diagnosed at later stages (3 or 4). While most patients are diagnosed following symptomatic presentation, no studies have compared symptoms and physical examination signs at or prior to diagnosis from electronic health records (EHR) in the United States (US).

**Objective:** To identify symptoms and signs in patients prior to lung cancer diagnosis in EHR data.

**Study Design:** Case-control study.

**Methods:** We studied 698 primary lung cancer cases in adults diagnosed between January 1, 2012 and December 31, 2019, and 6,841 controls matched by age, sex, smoking status, and type of clinic. Coded and free-text data from the EHR were extracted from 2 years prior to diagnosis date for cases and index date for controls. Univariate and multivariate conditional logistic regression were used to identify symptoms and signs associated with lung cancer. Analyses were repeated excluding symptom data from 1, 3, 6, and 12 months before the diagnosis/index dates.

**Results:** Eleven symptoms and signs recorded during the study period were associated with a significantly higher chance of being a lung cancer case in multivariate analyses. Of these, seven were significantly associated with lung cancer six months prior to diagnosis: hemoptysis (OR 3.2, 95%CI 1.9-5.3), cough (OR 3.1, 95%CI 2.4-4.0), chest crackles or wheeze (OR 3.1, 95%CI 2.3-4.1), bone pain (OR 2.7, 95%CI 2.1-3.6), back pain (OR 2.5, 95%CI 1.9-3.2), weight loss (OR 2.1, 95%CI 1.5-2.8) and fatigue (OR 1.6, 95%CI 1.3-2.1).

**Conclusions:** Patients diagnosed with lung cancer appear to have symptoms and signs recorded in the EHR that distinguish them from similar matched patients in ambulatory care, often six months or more before their diagnosis. These findings suggest opportunities to improve the diagnostic process for lung cancer in the US.

## Introduction

Lung cancer is the third most common cancer and the leading cause of cancer death in the United States (US).^1^ Most patients with lung cancer are diagnosed following presentation to healthcare settings with symptoms or diagnosed incidentally, and many patients (47%) present with late-stage disease (stages 3 or 4).^2^ Screening for lung cancer remains low in the US.^3,4^ In addition to optimizing screening, early detection efforts have focused on recognition of lung cancer symptoms with an overall goal of identifying patients at earlier, more treatable stages of the disease.^5–7^ These symptoms range from ‘alarm’ symptoms, such as hemoptysis (a rare symptom), to relatively non-specific symptoms, such as persistent cough or unexpected weight loss.^6^

Diagnosing lung cancer based on non-specific symptom presentation is challenging, as these symptoms are more commonly associated with benign conditions or may be overlooked for long periods of time. A study of over 43 million patients using Medicare claims data identified a median time from symptom onset to diagnosis of approximately six months.^8^ However, claims data lack the granularity needed to identify which clinical features patients present and how these might be used to differentiate patients with lung cancer from the vast majority of patients with benign conditions. To fill this gap, we examined the frequency and association of symptoms and physical examination signs in patients in ambulatory care prior to lung cancer diagnosis and matched controls.

## Methods

### Study design

We performed a case-control study using data from the University of Washington Medicine (UWM) electronic health records (EHR) and the Seattle/Puget Sound Surveillance, Epidemiology, and End Results (SEER) Program, a National Cancer Institute-supported national cancer registry. This study was approved by the University of Washington Human Subjects Division (STUDY 000013191).

### Setting

Cases and controls were identified from patients who received ambulatory care at UWM, a large tertiary care academic health center.

### Participants

Cases were identified from UWM patients aged 18 years or older, with a first primary lung cancer diagnosis (see International Classification of Diseases (ICD) 9 and 10 codes in e-Appendix 1) between January 1, 2012 and December 31, 2019, who had an established relationship with a UWM ambulatory care setting in the 2 years before the date of their first recorded lung cancer ICD code in the EHR (EHR diagnosis date). We chose the above study period because of the limited quality of the UWM EHR data prior to 2012. We defined ambulatory care as at least one encounter in family medicine, internal medicine, women’s health, obstetrics and gynecology, urgent care, and/or emergency medicine. We used linkage to the regional SEER registry to verify cancer incident cases. Cases were excluded if they did not match with the SEER registry or had evidence of a history of any of the following cancers identified using histology codes in SEER: tracheal cancer, mesothelioma, Kaposi sarcoma, lymphoma, or leukemia. Controls were identified from UWM patients with at least one encounter with the same type of ambulatory clinic within 3 months of the EHR diagnosis date of the index case (matching date). For each case, 10 controls were individually matched to the index case by age, sex (male, female), smoking status (ever vs. never), and type of ambulatory care clinic where lung cancer case presented (emergency medicine vs other clinics listed above). We chose a 10:1 control: case match because we recognize the wide variety of patients presenting to ambulatory care settings. Controls were excluded if they had any lung cancer ICD codes in their EHR prior to their matched case diagnosis (index) date. Excluded cancers in cases (based on histology codes from the SEER registry) were not identified in controls as registry data was not available for controls. We also excluded any cases and controls who did not have any ICD codes in any encounter in the 2 years prior to diagnosis date (cases) or index date (controls) to ensure availability of data on pre-diagnosis symptoms and signs.

### Data Collection

The UWM enterprise-wide data warehouse (EDW) was used to obtain data; this provides a central repository that integrates EHR across the UWM health care system including ambulatory care, specialty care and hospital services. Cases were identified during the study period using ICD codes (e-Appendix 1) and were linked to SEER to ensure accuracy of case identification and obtain history of previous cancers, histology (for exclusions and lung cancer type), and stage at diagnosis. The date of diagnosis was determined by date of pathology report at UWM. For cases that did not have a diagnosis through pathology or had a discrepancy greater than 30 days between date of pathology and first recorded lung cancer ICD code, two of three clinicians (MT, LKF, MAlA) reviewed the EHR of these cases to adjudicate dates. Controls were randomly sampled from within the matching strata, based on this adjudicated date of diagnosis.

Cases who had undergone lung cancer screening using low-dose computed tomography (LDCT) within the 12 months prior to diagnosis date were identified from billing code (Current Procedural Terminology or CPT 71271) and/or ICD codes (V76.0 [ICD-9] or Z12.2 [ICD-10].

An EHR data extraction protocol was applied to all encounters in the 2-year period prior and up to six months following the diagnosis date (cases) and index date (controls). These data comprised of demographics (e.g., age, sex, race, ethnicity), all ICD codes and CPT procedure codes linked to encounters such as laboratory tests, imaging procedures, and pathology data. We also extracted corresponding unstructured clinical notes for any of the above encounters. ICD codes recorded during the 2-year period prior to diagnosis for cases or prior to index date for controls were searched for the presence of 31 potential comorbidities to calculate the Elixhauser comorbidity index.^9^ We excluded lung cancer ICD code information from this calculation. These index scores were then used to calculate van Walraven weighted scores for each patient, a range of -19 to 89.^10,11^

### Symptoms and signs

We identified symptoms and signs using coded data and unstructured data. A list of symptoms and signs which have previously been reported in cohort or case-control studies of individuals with lung cancer were identified from systematic reviews, hand review of individual studies, and from contact with experts in oncology, cardiothoracic surgery, and primary care (FW, RN, FF, MT, see e-Appendix 2).^5,6,12–17^ These were mapped to ICD codes, and used to search the extracted EHR coded data for any encounters that included any of these ICD codes in the 2-year observation period.

Symptoms and signs were automatically extracted from free-text clinical notes using natural language processing (NLP), including notes for all visit types in the 2-year period. In previous work, we developed a deep learning symptom extraction model using the COVID-19 Annotated Clinical Text Corpus (CACT),^18^ which was then adapted to the lung cancer domain. This involved creating the Lung Cancer Annotated Clinical Text (LACT) Corpus, composed of 270 notes from lung cancer patients (170 training and 100 test notes).^19^ We trained the lung cancer symptom extractor by combining the CACT and LACT training sets. On the LACT test set, the lung cancer symptom extractor achieved 0.72 F1 for symptom identification and 0.65 F1 for assertion prediction. This extraction performance is comparable to the LACT inter-rater agreement of 0.82 F1 for symptom identification and 0.79 F1 for assertion prediction, indicating the model is achieving approximately human-level performance. We included the extracted symptoms and signs with assertion value present.

### Data analysis

Frequencies and counts were calculated for characteristics of cases and controls. The number of symptoms and signs obtained from coded data was compared to that obtained from free-text data using descriptive statistics. The proportion of patients with evidence of each symptom/sign occurring in the 2-year period prior to the diagnosis or index date was described for cases and controls. Odds of patients’ case status, based on symptoms and signs identified from a combined dataset of coded and free-text data, were estimated using unadjusted conditional logistic regression. Symptoms and signs associated with lung cancer in unadjusted regressions (*p* < 0.1) were included into multivariate conditional logistic regression analyses. We used the van Walraven comorbidity score to adjust for population differences in comorbidity burden. Analyses were repeated excluding symptom and sign data from 1, 3, 6, and 12 months before the diagnosis (or index) date. Lag times were chosen to provide information on the pattern of symptom-related visits over time and identify the symptoms and signs presenting furthest from diagnosis. We conducted secondary analyses investigating the potential effect of chronic respiratory disease (CRD) status, as defined by the presence of ICD codes within the Elixhauser chronic respiratory disease subgroup, on presence of symptoms and signs in the pre-diagnostic interval. We expected patients with CRD to present with symptoms and signs similar to those that present in early lung cancer. We assessed the effect of CRD by repeating the conditional logistic regression model including CRD as a covariate.

Statistical analyses were conducted using Python 3.7 with the packages SciPy (version 1.4.1) and Statsmodels (version 0.11.1). The study was reported in line with the STROBE guidelines.^20^

## Results

### Participants

#### Selection of cases & controls

A total of 7,883 patients with lung cancer ICD codes were identified in the UWM EDW over the study period. Following linkage of these patients and those identified as having a primary lung tumor from SEER, 4,115 patients were identified common to both, including 741 cases. After matching 7,410 controls, a chart review resulted in exclusion of 43 additional cases. Controls that were matched to these 43 cases were excluded (n = 422), resulting in 698 cases matched to 6,841 controls.

### Description of cases and controls

Cases and controls were similar in terms of sex and race (cases 50.6% male, 75.5% White; controls 50.5% male, 75.7% White, see Table 1). Cases had higher comorbidity scores (*M* = 14.9, *SD* = 11.6) than controls (*M* = 4.4, *SD =* 8.6). Cases also had a greater median number of health care visits over the 2-year period prior to diagnosis (51.0, 95%CI: 28.0-97.8) than controls (23.0, 95%CI: 9.0-53.0). The difference in median number of health care visits was greater in the last 3-month period prior to the diagnosis/index date (cases 21.0, 95%CI: 12.0-35.0 vs. controls 5.0, 95%CI: 2.0-11.0) than in the 2^nd^, 3^rd^, or 4^th^ quarters prior to diagnosis. The stage distribution of cases was as follows: Stage 1-29%, Stage 2-7%, Stage 3-17%, and Stage 4 -42% (5% were Stage 0 or Unknown Stage).

**Table 1.**
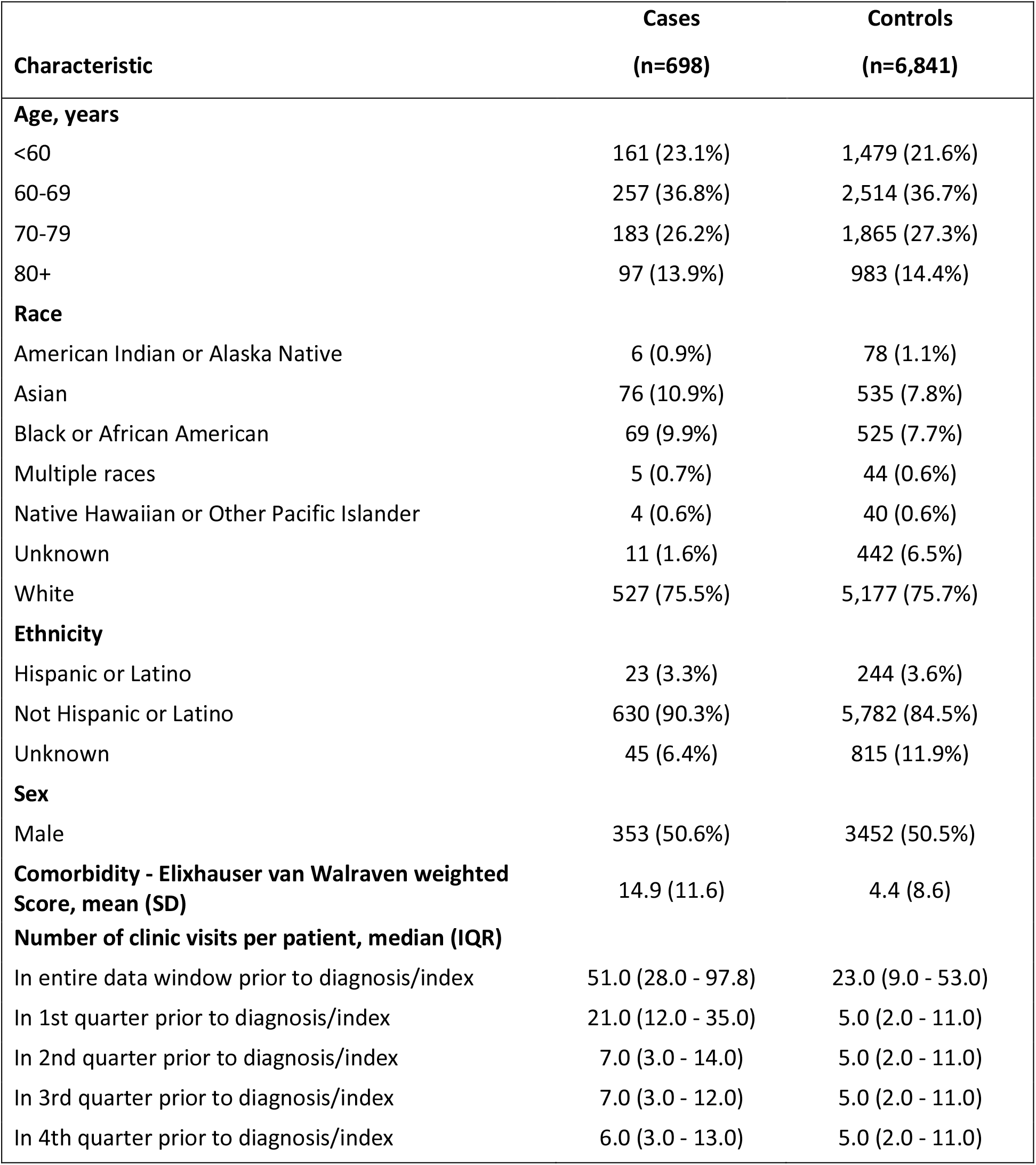
Characteristics of patients with lung cancer (cases) and matched controls in ambulatory care.

| **Characteristic** | **Cases**<br>**(n=698)** | **Controls**<br>**(n=6,841)** |
| --- | --- | --- |
| **Age, years** |  |  |
| <60 | 161 (23.1%) | 1,479 (21.6%) |
| 60-69 | 257 (36.8%) | 2,514 (36.7%) |
| 70-79 | 183 (26.2%) | 1,865 (27.3%) |
| 80+ | 97 (13.9%) | 983 (14.4%) |
| **Race** |  |  |
| American Indian or Alaska Native | 6 (0.9%) | 78 (1.1%) |
| Asian | 76 (10.9%) | 535 (7.8%) |
| Black or African American | 69 (9.9%) | 525 (7.7%) |
| Multiple races | 5 (0.7%) | 44 (0.6%) |
| Native Hawaiian or Other Pacific Islander | 4 (0.6%) | 40 (0.6%) |
| Unknown | 11 (1.6%) | 442 (6.5%) |
| White | 527 (75.5%) | 5,177 (75.7%) |
| **Ethnicity** |  |  |
| Hispanic or Latino | 23 (3.3%) | 244 (3.6%) |
| Not Hispanic or Latino | 630 (90.3%) | 5,782 (84.5%) |
| Unknown | 45 (6.4%) | 815 (11.9%) |
| **Sex** |  |  |
| Male | 353 (50.6%) | 3452 (50.5%) |
| **Comorbidity - Elixhauser van Walraven weighted Score, mean (SD)** | 14.9 (11.6) | 4.4 (8.6) |
| **Number of clinic visits per patient, median (IQR)** |  |  |
| In entire data window prior to diagnosis/index | 51.0 (28.0 - 97.8) | 23.0 (9.0 - 53.0) |
| In 1st quarter prior to diagnosis/index | 21.0 (12.0 - 35.0) | 5.0 (2.0 - 11.0) |
| In 2nd quarter prior to diagnosis/index | 7.0 (3.0 - 14.0) | 5.0 (2.0 - 11.0) |
| In 3rd quarter prior to diagnosis/index | 7.0 (3.0 - 12.0) | 5.0 (2.0 - 11.0) |
| In 4th quarter prior to diagnosis/index | 6.0 (3.0 - 13.0) | 5.0 (2.0 - 11.0) |

### Frequency of symptoms and signs extracted from coded and free-text data

Of the 22 symptoms and signs that we systematically examined, NLP identified 20 of the 22 symptoms and signs in greater proportions of patients affected than from the coded data alone (see e-Appendix 3). In comparison to coded data, we saw a range of 12.9% to 97.6% greater symptom and signs reports with NLP of textual clinical notes. In contrast, a greater proportion of patients had two symptoms and signs (shoulder pain, lymphadenopathy) identified from coded rather than free-text data.

### Comparison of frequency of symptoms and signs between cases and controls

The frequency of all 22 symptoms and signs examined was higher in cases than controls (see Table 2). Moreover, the ranking of symptoms and signs differed slightly between cases and controls, with cases reporting cough (82.1%), shortness of breath (73.8%), fatigue (68.2%), ankle swelling (64.0%), and chest pain (57.7%), whereas controls reported ankle swelling (26.9%), cough (24.2%), shortness of breath (23.6%), fatigue (23.2%) and chest pain (20.5%) most frequently. Hemoptysis occurred relatively infrequently among cases (16.5%) and rarely among controls (1.0%).

**Table 2.** Comparison of frequency of symptoms and signs identified in coded or free-text data in cases compared to controls.

| **Symptom or sign** | **Cases**<br>**(n=698)** | **Controls**<br>**(n=6,841)** |
| --- | --- | --- |
| Cough | 573 (82.1%) | 1,654 (24.2%) |
| Shortness of breath | 515 (73.8%) | 1,613 (23.6%) |
| Fatigue | 476 (68.2%) | 1,587 (23.2%) |
| Ankle swelling | 447 (64.0%) | 1,838 (26.9%) |
| Chest Pain | 403 (57.7%) | 1,401 (20.5%) |
| Chest crackles or wheeze | 397 (56.9%) | 575 (8.4%) |
| Back pain | 350 (50.1%) | 946 (13.8%) |
| Change in bowel habits | 336 (48.1%) | 1,155 (16.9%) |
| Muscle weakness | 334 (47.9%) | 1,102 (16.1%) |
| Fever | 322 (46.1%) | 1,334 (19.5%) |
| Weight loss | 308 (44.1%) | 522 (7.6%) |
| Headache | 304 (43.6%) | 1,205 (17.6%) |
| Dizziness | 299 (42.8%) | 1,319 (19.3%) |
| Bone pain | 270 (38.7%) | 725 (10.6%) |
| Lack of appetite | 196 (28.1%) | 457 (6.7%) |
| Shoulder pain | 180 (25.8%) | 713 (10.4%) |
| Lymphadenopathy | 151 (21.6%) | 105 (1.5%) |
| Night sweats | 150 (21.5%) | 371 (5.4%) |
| Changes in sleep | 134 (19.2%) | 631 (9.2%) |
| Hemoptysis | 115 (16.5%) | 67 (1.0%) |
| Hoarseness | 67 (9.6%) | 133 (1.9%) |
| Finger clubbing | 39 (5.6%) | 2 (0.0%) |

### Univariate associations of symptoms and signs between cases and controls

In models adjusted for comorbidity score, when considered independently, all 22 symptoms and signs had odds ratios that were significantly different between cases and controls (all *p* < 0.0001, see Table 3). The symptoms and signs with the largest odds ratios (OR) significantly associated with a higher chance of being a case were finger clubbing (OR 175.7, 95%CI: 40.1-770.0), hemoptysis (OR 14.5, 95%CI: 10.2-20.8), cough (OR 11.1, 95%CI: 8.8-13.9), chest crackles or wheeze (OR 9.9, 95%CI: 8.1-12.2), and lympadenopathy (OR 9.4, 95%CI: 6.9-12.8).

**Table 3.** Univariate and multivariate analyses of symptoms and signs identified in coded or free-text data of cases compared to controls, adjusted for comorbidity (descending order by multivariate odds ratios)

| **Symptom or sign** | **Univariate**<br>**Odds ratio (95%CI)** | **Multivariate**<br>**Odds ratio (95%CI)** | **Multivariate**<br>**P value** |
| --- | --- | --- | --- |
| Finger clubbing | 175.7 (40.1 - 770.0)* | 50.1 (8.9 - 283.3) | <0.0001 |
| Lymphadenopathy | 9.4 (6.9 - 12.8)* | 5.8 (3.8 - 8.8) | <0.0001 |
| Cough | 11.1 (8.8 - 13.9)* | 4.7 (3.5 - 6.3) | <0.0001 |
| Hemoptysis | 14.5 (10.2 - 20.8)* | 3.5 (2.2 - 5.5) | <0.0001 |
| Chest crackles or wheeze | 9.9 (8.1 - 12.2)* | 3.2 (2.4 - 4.3) | <0.0001 |
| Weight loss | 5.9 (4.8 - 7.2)* | 2.9 (2.2 - 3.9) | <0.0001 |
| Back pain | 4.7 (3.9 - 5.7)* | 2.4 (1.8 - 3.1) | <0.0001 |
| Bone pain | 4.6 (3.8 - 5.7)* | 2.3 (1.7 - 3.1) | <0.0001 |
| Shortness of breath | 6.0 (4.9 - 7.3)* | 1.9 (1.4 - 2.5) | <0.0001 |
| Fatigue | 4.8 (4.0 - 5.8)* | 1.8 (1.4 - 2.4) | <0.0001 |
| Chest Pain | 3.6 (3.0 - 4.3)* | 1.4 (1.1 - 1.8) | 0.0118 |
| Shoulder pain | 2.3 (1.8 - 2.8)* | 1.3 (1.0 - 1.7) | 0.1111 |
| Ankle swelling | 3.3 (2.7 - 4.0)* | 1.1 (0.9 - 1.5) | 0.3643 |
| Headache | 2.5 (2.1 - 3.0)* | 1.1 (0.8 - 1.4) | 0.5619 |
| Hoarseness | 3.5 (2.5 - 5.0)* | 1.1 (0.7 - 1.7) | 0.8447 |
| Change in bowel habits | 3.0 (2.5 - 3.6)* | 1.0 (0.8 - 1.4) | 0.8880 |
| Muscle weakness | 2.9 (2.4 - 3.5)* | 1.0 (0.7 - 1.3) | 0.9581 |
| Night sweats | 3.3 (2.6 - 4.2)* | 0.8 (0.6 - 1.2) | 0.2998 |
| Lack of appetite | 2.6 (2.1 - 3.3)* | 0.7 (0.5 - 0.9) | 0.0193 |
| Dizziness | 2.0 (1.7 - 2.4)* | 0.6 (0.4 - 0.8) | 0.0004 |
| Changes in sleep | 1.3 (1.1 - 1.7)* | 0.5 (0.3 - 0.6) | <0.0001 |
| Fever | 2.1 (1.7 - 2.5)* | 0.4 (0.3 - 0.6) | <0.0001 |

*Note:* Conditional logistic regression models adjusted for comorbidities using van Walraven weighted score with each symptom or sign modeled individually (univariate) and mutually adjusted (multivariate)
\*Significant at $p < 0.0001$ for univariate analysis

### Multivariable associations of symptoms and signs between cases and controls

We included all 22 symptoms and signs from the univariate analysis and comorbidity score in a multivariate analysis. After mutual adjustment, 15 had significant ORs (all *p* < 0.05, see Table 3). The presence of 11 symptoms and signs were associated with a significantly higher odds of being a case, with ORs ranging from 1.4 (chest pain) to 50.1 (finger clubbing). The largest ORs were noted for finger clubbing (OR 50.1, 95%CI: 8.9-283.3), lymphadenopathy (OR 5.8, 95%CI: 3.8-8.8), cough (OR 4.7, 95%CI: 3.5-6.3), hemoptysis (OR 3.5, 95%CI: 2.2-5.5) and chest crackles or wheeze (OR 3.2, 95%CI: 2.4-4.3). In contrast, the presence of four symptoms was associated with a significantly higher odds of being a control: fever (OR 0.4, 95%CI: 0.3-0.6), changes in sleep (OR 0.5, 95%CI: 0.3-0.6), dizziness (OR 0.6, 95%CI: 0.4-0.8), and lack of appetite (OR 0.7, 95%CI: 0.5-0.9).

We repeated the multivariate analysis, excluding symptoms and signs recorded in periods of 1, 3, 6 and 12 months prior to diagnosis (see Figure 2). Some symptoms and signs remained significantly associated with cases up to 6 months prior to diagnosis (cough, hemoptysis, chest crackles and wheeze, weight loss, back pain, bone pain, fatigue). Of these, all except weight loss were also significantly associated with cases 12 months prior to diagnosis. Other symptoms and signs became significantly associated with being a case closer to the date of diagnosis: shortness of breath and chest pain (3 months prior to diagnosis), lymphadenopathy and finger clubbing (1 month prior) (see e-Appendix 4).

**Figure 1.**
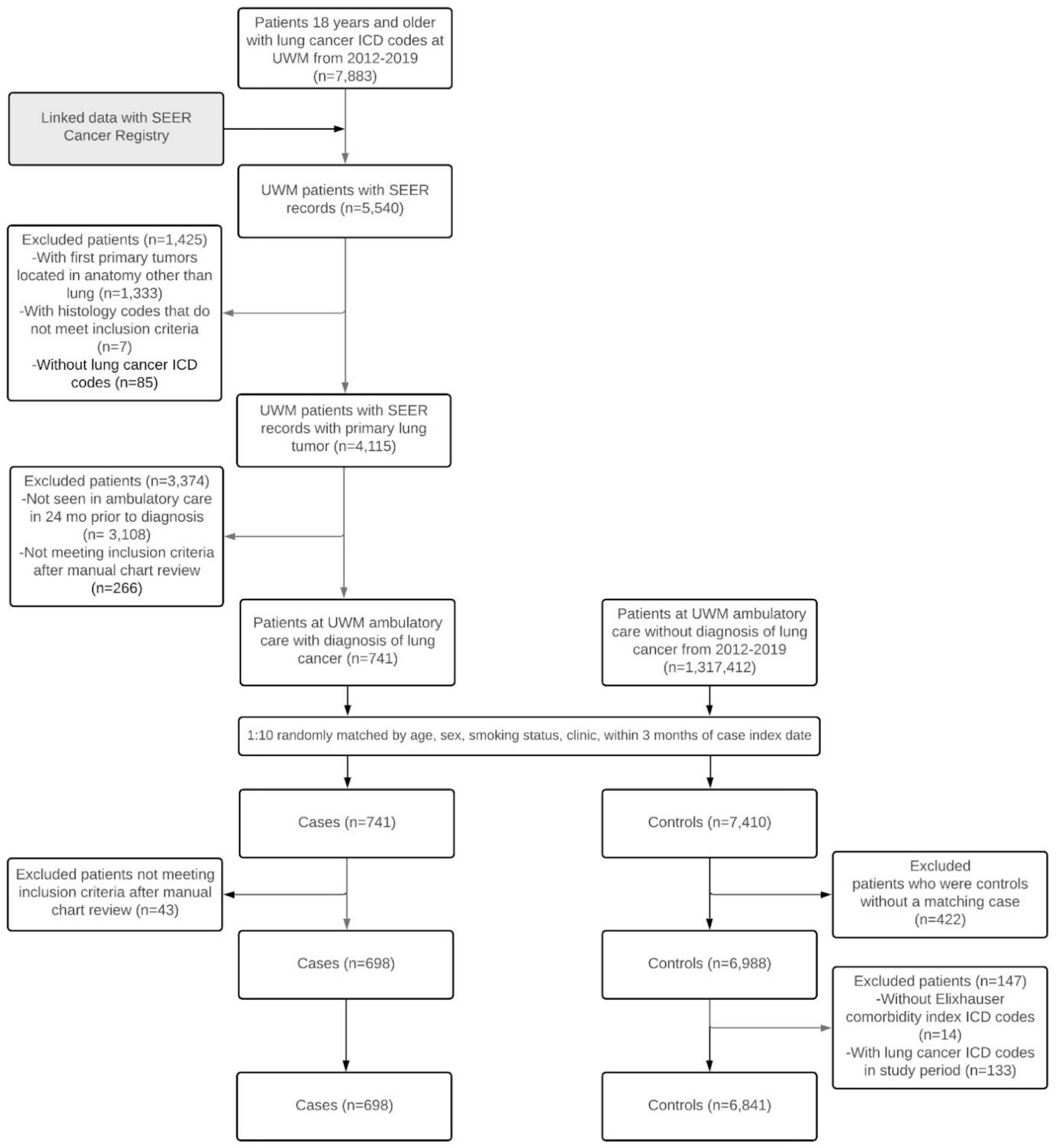
Flow chart of case and control selection.

**Figure 2:**
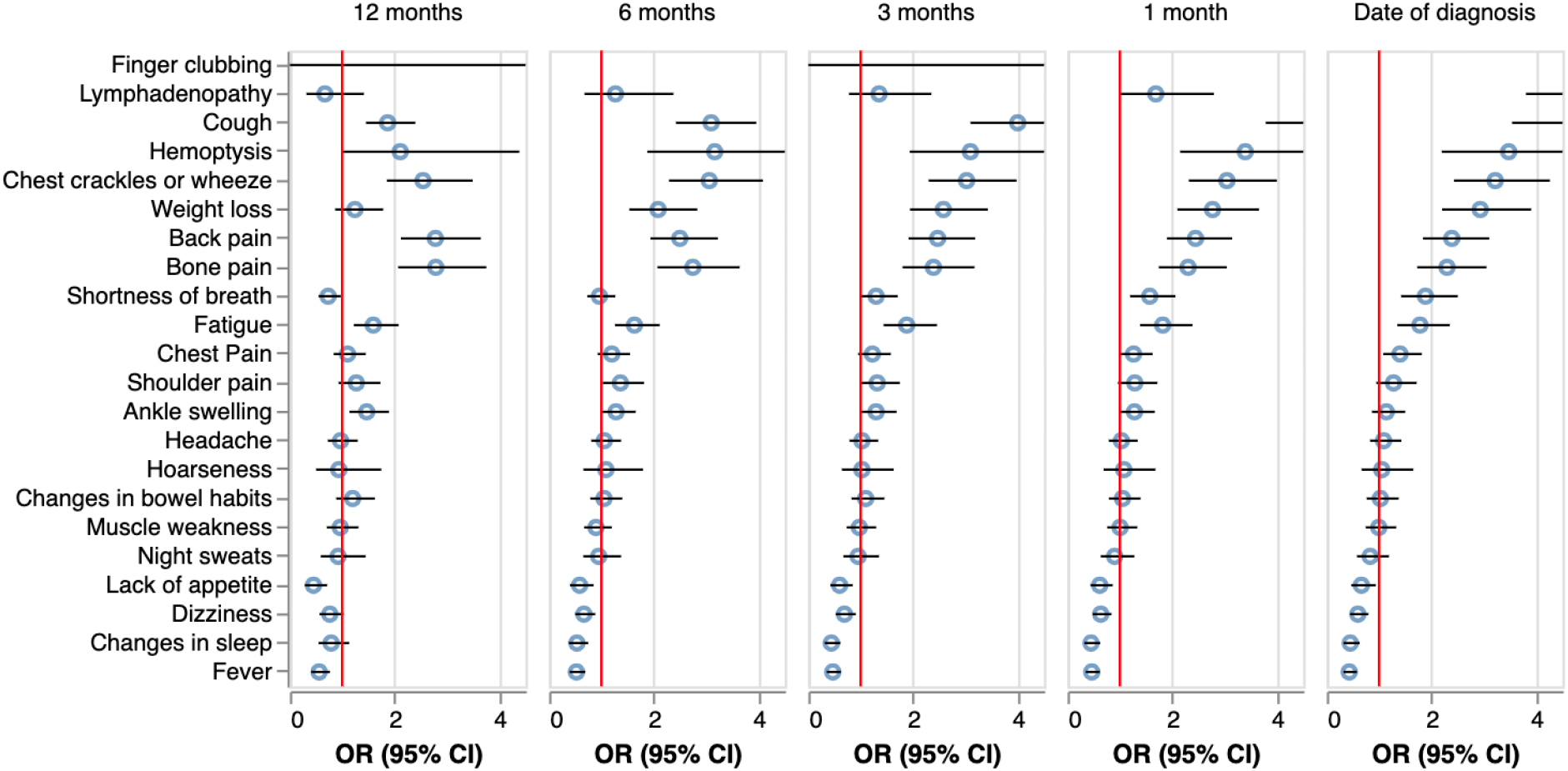
Multivariable analysis of symptoms or signs of cases compared to controls with symptom and sign data excluded from 1, 3, 6, and 12 months prior to diagnosis/index date. *Note*: Mutual adjustment of all symptoms and signs in using a conditional logistic regression model stratified by time prior to date of diagnosis. Models additionally adjusted for comorbidities using van Walraven weighted score.

### Secondary analyses

To determine whether the associations were robust to the presence of CRD, we performed a secondary conditional logistic regression that was adjusted for CRD, along with all our matching variables and comorbidity score. The presence of CRD appeared to have no statistically significant effect when directly added as a covariate (OR: 1.05, 95%CI: (0.81, 1.36, *p* = 0.7229, see Appendices 5 & 6).

## Discussion

### Main findings

This is the first case-control study in the US to use routine, prospectively collected EHR data to describe the frequency of symptoms and signs of lung cancer and estimate associations with incident lung cancer cases compared to non-lung cancer patients receiving routine ambulatory care in the same time period. Our findings provide unique information on symptoms and signs associated with a higher chance of a patient in ambulatory care being diagnosed with lung cancer, and the duration of these associations prior to their cancer diagnosis. In contrast to prior work on national databases, extracting clinicians’ documentation of clinical features from their free text clinical notes using NLP provided more complete symptom identification data, rather than relying on data available only in coded, structured data collected in routine care. Our findings provide evidence-based, quantitative support for the development of decision rules around the diagnostic workup of symptomatic patients, which could lead to the improvement of earlier diagnosis of lung cancer. Of the 22 symptoms and signs studied, 11 were found in adjusted models to be associated with a higher chance of being a lung cancer case, and most of these 11 were present and still significantly associated up to 12 months prior to diagnosis; this suggests opportunities for improved screening practices that may lead to earlier diagnosis and possibly improved outcomes.

Our findings also suggest that the clinical presentation of lung cancer appears to be similar, regardless of the presence of other comorbidities, CRD, or smoking. For patients and clinicians this is important as several of the symptoms or signs we identified may currently be dismissed as being attributable to underlying smoking or comorbid conditions.

### Comparison with existing literature

Several of the symptoms and signs we found as having statistically significant odds ratios have been identified in studies using data from ambulatory care in other healthcare systems, especially hemoptysis and cough. However, among the symptoms and signs Hamilton and colleagues (2005) found to be associated with being a lung cancer case in the United Kingdom (UK), loss of appetite had the highest OR (86.0), whereas we failed to identify an association with lung cancer.^5^ This may be due to a difference in study populations or our use of NLP in EHR data.

Our findings also provide evidence of the temporality of a ‘clinical signal’ for lung cancer based on symptoms and signs documented in the EHR, at least six and up to 12 months prior to diagnosis, consistent with a Medicare claims study. Data from our study and Nadpara and colleagues’ (2015) study, which used claims data, provide evidence for time intervals from first presentation with symptoms to diagnosis that are on the upper range (six months) of those reported using analysis of coded symptoms in primary care databases in several UK and European studies.^8^ These describe the overall time interval from first symptom recording in medical records to diagnosis ranging from 3- to 6-months.^6,21,22^ While not directly comparable, qualitative research from patients with lung cancer and caregivers describe changes noticeable to the individual more than 12 months before attending a health care visit.^16,23,24^

### Strengths and limitations

Using NLP to extract symptoms and signs from unstructured data allowed us to capture a more complete dataset of symptom presence compared to using coded data alone. We selected cases from an empaneled ambulatory care population, where we expected EHR data would be available for the period of interest in this study and attempted to exclude patients who were attending only for secondary or tertiary care provided at UWM. Controls were randomly selected based on case clinic type, to reduce the possibility of bias, and duration of follow-up time and availability of data for cases and controls were similar, particularly in visit frequency. We used a robust design where we matched 10 controls to 1 case, providing greater power and precision, and matched on smoking so that our analyses could not be confounded based on ever vs. never exposure to smoking.

Limitations included criteria for selection of cases and controls differed slightly. As is customary in incident case-control studies, cases were selected based on a diagnosis date defined as the date of the first lung cancer ICD code in the EHR. In this way, we captured the diagnostic path from symptom presentation to diagnosis for all cases. Controls were selected based on having a visit to the matched case clinic type (to account for difference in emergency vs other forms of ambulatory care) within 3 months of the case diagnosis date (to avoid potential seasonal differences in respiratory symptoms), however the timing of control selection does not necessarily reflect a “pathway to diagnosis” for some other condition, just recent routine care. Additionally, because we did not link to SEER for the control population, we were unable to apply two of the case exclusion criteria to our control sample: no current or prior history of lung cancer in SEER, although we did check the UW EHR for concurrent lung-cancer related ICD codes and medical history so this should be rare, and no prior history of tracheal cancer, mesothelioma, Kaposi sarcoma, lymphoma, or leukemia in SEER. Additionally, EHR data can sometimes be subject to misclassification. For example, detailed EHR smoking history may be unreliable and the EHR does not reliably capture health literacy or socioeconomic status; however, we used a very broad definition of smoking (ever vs. never) and used a comorbidity score to control for health status. Finally, availability and timing of symptom data for cases and controls is based on patient interactions with the healthcare system, not a pre-specified protocol of data collection. Patients who have more contact with their providers (which could be due to a range of factors) may have had more data captured.

### Implications for clinicians, researchers, policy makers

Differentiating patients who may have symptoms or signs of lung cancer from those attending ambulatory care is a critical and challenging step in the earlier detection of this cancer. Our findings not only identify the ‘red flag’ (highly specific, but infrequent) symptoms and signs that primary care providers should be aware of (e.g., hemoptysis), but also highlight which of a larger range of ‘non-specific’ symptoms and signs should equally raise suspicion such as bone pain and weight loss. Furthermore, our findings support the importance of clinical documentation, and continuity of care to identify and act on sustained changes in patients’ clinical presentations.

Confirmation of our findings using datasets from other healthcare systems in the U.S. are needed and could be enhanced by more advanced machine learning modelling to incorporate additional clinical variable including quantitative data such as changes in body weight or results of routinely collected laboratory tests, given emerging evidence for associations between weight loss and minor deviations of hemoglobin or platelet count with incident cancer.^25^ Given the low uptake of low dose CT screening for lung cancer in the U.S., our findings provide support for revising current priorities to improve early diagnosis of lung cancer.^26^

### Conclusions

Patients in ambulatory care settings who are subsequently diagnosed with lung cancer appear to have symptoms and signs that distinguish them from other patients, often months before lung cancer diagnosis. To improve earlier detection of lung cancer, interventions are urgently needed that promote earlier screening based on symptomatic presentations in ambulatory care that may lead to an earlier detection and treatment of lung cancer.

## Data Availability

All data produced in the present study are available upon reasonable request to the authors.

## Abbreviation List

CACT: COVID-19 Annotated Clinical Text Corpus
CPT: Current Procedural Terminology
CRD: Chronic respiratory disease
EDW: Enterprise-wide data warehouse
EHR: Electronic health records
ICD: International Classification of Diseases
LACT: Lung Cancer Annotated Clinical Text Corpus
LDCT: Low-dose computed tomography
NLP: Natural language processing
SEER: Seattle/Puget Sound Surveillance, Epidemiology, and End Results
UWM: University of Washington Medicine

## Acknowledgments

This research was funded by the Gordon and Betty Moore Foundation (GBMF8837) and the CanTest Collaborative, funded by Cancer Research UK (RG85791). This research was supported by the Cancer Surveillance System of the Fred Hutchinson Cancer Research Center, which is funded by Contract No. May 2018 – April 2028: HHSN261201800004I; NCI Control Number: N01 PC-2018-00004 from the Surveillance, Epidemiology and End Results (SEER) Program of the National Cancer Institute with additional support from the Fred Hutchinson Cancer Research Center and the State of Washington.

## Notes

**Conflicts of interest:** The authors have no conflicts of interest to declare.

### Competing Interest Statement

The authors have declared no competing interest.

### Author Declarations

The Human Subjects Division/IRB of the University of Washington gave ethical approval for this work (STUDY 000013191).

