## Appendices for "Symptoms and signs of lung cancer prior to diagnosis: Comparative study using electronic health records"

### Appendix 1. Diagnostic codes used to identify cases of lung cancer

ICD 9: 162.2 – 162.9

- 162.2 - Malignant neoplasm of main bronchus
- 162.3 - Malignant neoplasm of upper lobe, bronchus or lung
- 162.4 - Malignant neoplasm of middle lobe, bronchus or lung
- 162.5 - Malignant neoplasm of lower lobe, bronchus or lung
- 162.8 - Malignant neoplasm of other parts of bronchus or lung
- 162.9 - Malignant neoplasm of bronchus and lung, unspecified

ICD 10: C34.0 – C34.9

- C34.0 - Malignant neoplasm of main bronchus
- C34.00 - Malignant neoplasm of unspecified main bronchus
- C34.01 - Malignant neoplasm of right main bronchus
- C34.02 - Malignant neoplasm of left main bronchus
- C34.1 - Malignant neoplasm of upper lobe, bronchus or lung
- C34.10 - Malignant neoplasm of upper lobe, unspecified bronchus or lung
- C34.11 - Malignant neoplasm of upper lobe, right bronchus or lung
- C34.12 - Malignant neoplasm of upper lobe, left bronchus or lung
- C34.2 - Malignant neoplasm of middle lobe, bronchus or lung
- C34.3 - Malignant neoplasm of lower lobe, bronchus or lung
- C34.30 - Malignant neoplasm of lower lobe, unspecified bronchus or lung
- C34.31 - Malignant neoplasm of lower lobe, right bronchus or lung
- C34.32 - Malignant neoplasm of lower lobe, left bronchus or lung
- C34.8 - Malignant neoplasm of overlapping sites of bronchus and lung
- C34.80 - Malignant neoplasm of overlapping sites of unspecified bronchus and lung
- C34.81 - Malignant neoplasm of overlapping sites of right bronchus and lung
- C34.82 - Malignant neoplasm of overlapping sites of left bronchus and lung
- C34.9 - Malignant neoplasm of unspecified part of bronchus or lung
- C34.90 - Malignant neoplasm of unspecified part of unspecified bronchus or lung
- C34.91 - Malignant neoplasm of unspecified part of right bronchus or lung
- C34.92 - Malignant neoplasm of unspecified part of left bronchus or lung

Excluded ICD Diagnostic Codes

- ICD-9: 162.0
- ICD-10: C33

Excluded Histology codes

- Mesothelioma: 9050-9055
- Kaposi Sarcoma: 9140
- Lymphoma/leukemia: M9590-M9992

**Appendix 2. Symptoms and signs Identified in peer-reviewed literature previously associated with lung cancer in primary care populations**

| Symptom or sign | ICD 9 code(s) | ICD10 code(s) | References |
|---|---|---|---|
| Ankle swelling | 782.3 | R60.9 | [1]Ellis (2011) |
| Back pain | 724.1 | M54.6 | [1]Ellis (2011) [2]Molassiotis (2010) |
| Bone pain | 733.9 | M85.80 | [3]Gould (2008) [4]Nadpara (2015) |
| Changes in bowel habits | 787.99 | R19.4 | [5]Corner (2005) |
| Changes in sleep | 780.50 | G47.9 | [5]Corner (2005) |
| Chest Pain | 786.5<br>786.50<br>786.51<br>786.52<br>786.59 | R07.9<br>R07.81 | [1]Ellis (2011) [4]Nadpara (2015) [5]Corner (2005) [6]Chowienczyk, Hamilton (2020) [7]Walter (2015) [8]Hamilton (2005) [9]Ades (2014) [10]Redaniel (2015) [11]Tod (2008) [12]Mitchell (2013) |
| Chest crackles or wheeze | 786.7 | R09.89 | [10]Redaniel (2015) |
| Cough | 786.2<br>491.0 | R05 | [1]Ellis (2011) [2]Molassiotis (2010) [4]Nadpara (2015) [5]Corner (2005) [6]Chowienczyk, Hamilton (2020) [7]Walter (2015) [9]Ades (2014) [10]Redaniel (2015) [11]Tod (2008) [12]Mitchell (2013) [13]Menon (2019) |
| Dizziness | 780.4 | R42 | [2]Molassiotis (2010) |
| Fatigue/tiredness | 780.79 | R53.81<br>R53.8<br>R53.83<br>R53.1 | [1]Ellis (2011) [2]Molassiotis (2010) [5]Corner (2005) [6]Chowienczyk, Hamilton (2020) [7]Walter (2015) [8]Hamilton (2005) [10]Redaniel (2015) [11]Tod (2008) [13]Menon (2019) |
| Fever | 780.6<br>780.60 | R50.9 | [4]Nadpara (2015) |
| Finger clubbing | 781.5 | R68.3 | [4]Nadpara (2015) [8]Hamilton (2005) [10]Redaniel (2015) |
| Headache | 784.0 | R51 | [1]Ellis (2011) |
| Hemoptysis | 786.3<br>786.30<br>786.39 | R04.2 | [1]Ellis (2011) [4]Nadpara (2015) [5]Corner [6]Chowienczyk, Hamilton (2020) [7]Walter (2015) [8]Hamilton (2005) [10]Redaniel (2015) (2005) [11]Tod (2008) [12]Mitchell (2013) [13]Menon (2019) [14]Hippisley-Cox (2011) |

| | | | |
|---|---|---|---|
| Hoarseness | 784.49<br>784.42 | R49.8<br>R49.0 | [1]Ellis (2011) [2]Molassiotis (2010) [7]Walter (2015) [10]Redaniel (2015) [11]Tod (2008) [12]Mitchell (2013) |
| Lack of appetite | 783 | R63.0 | [1]Ellis (2011) [2]Molassiotis (2010) [5]Corner (2005) [6]Chowienczyk, Hamilton (2020) [7]Walter (2015) [8]Hamilton (2005) [13]Menon (2019) |
| Lympadenopathy | 785.6 | R59.9 | [10]Redaniel (2015) [12]Mitchell (2013) |
| Muscle weakness | 728.87 | M62.81 | [4]Nadpara (2015) [12]Mitchell (2013) |
| Night sweats | 780.8 | R61 | [3]Gould (2008) [5]Corner (2005) |
| Shortness of breath | 786.05<br>786.0<br>786.9 | R06.02<br>R06.00<br>R06.09 | [1]Ellis (2011) [2]Molassiotis (2010) [4]Nadpara (2015) [5]Corner (2005) [6]Chowienczyk, Hamilton (2020) [7]Walter (2015) [8]Hamilton (2005) [10]Redaniel (2015) [12]Mitchell (2013) [13]Menon (2019) |
| Shoulder pain | 719.41 | M25.511<br>M25.512<br>M25.519 | [10]Redaniel (2015) [12]Mitchell (2013) |
| Weight loss | 783.21 | R63.4 | [1]Ellis (2011) [4]Nadpara (2015) [5]Corner (2005) [6]Chowienczyk, Hamilton (2020) [7]Walter (2015) [8]Hamilton (2005) [10]Redaniel (2015) [11]Tod (2008) [12]Mitchell (2013) |
| Wheezing and stridor | 786.07<br>786.1 | R06.2<br>R06.1 | [4]Nadpara (2015) [10]Redaniel (2015) |

**Appendix 3. Comparison of the number of patients with symptoms and signs extracted from the electronic medical record of cases or controls from coded fields versus free-text data using natural language processing (NLP)**

| Symptom or sign | Identified from NLP (% of patients) | Identified from coded data (% of patients) | Identified from either coded data or NLP (% of patients) | NLP adds (NLP adds n/coded or NLP n) |
|---|---|---|---|---|
| Cough | 1700 (22.6%) | 1139 (15.1%) | 2227 (29.5%) | 1088 (48.9%) |
| Shortness of breath | 1580 (21.0%) | 1111 (14.7%) | 2128 (28.2%) | 1017 (47.8%) |
| Chest Pain | 1241 (16.5%) | 981 (13.0%) | 1804 (23.9%) | 823 (45.6%) |
| Fatigue | 1489 (19.8%) | 959 (12.7%) | 2063 (27.4%) | 1104 (53.5%) |
| Shoulder pain | 513 (6.8%) | 594 (7.9%) | 893 (11.9%) | 299 (33.5%) |
| Dizziness | 1331 (17.7%) | 536 (7.1%) | 1618 (21.5%) | 1082 (66.9%) |
| Ankle swelling | 2081 (27.6%) | 509 (6.8%) | 2285 (30.3%) | 1776 (77.7%) |
| Headache | 1281 (17.0%) | 415 (5.5%) | 1509 (20.0%) | 1094 (72.5%) |
| Weight loss | 646 (8.6%) | 328 (4.4%) | 830 (11.0%) | 502 (60.5%) |
| Fever | 1517 (20.1%) | 252 (3.3%) | 1656 (22.0%) | 1404 (84.8%) |
| Chest crackles or wheeze | 834 (11.1%) | 242 (3.2%) | 972 (12.9%) | 730 (75.1%) |
| Lympadenopathy | 52 (0.7%) | 223 (3.0%) | 256 (3.4%) | 33 (12.9%) |
| Bone pain | 829 (11.0%) | 216 (2.9%) | 995 (13.2%) | 779 (78.3%) |
| Muscle weakness | 1327 (17.6%) | 205 (2.7%) | 1436 (19.1%) | 1231 (85.7%) |
| Back pain | 1220 (16.2%) | 154 (2.0%) | 1296 (17.2%) | 1142 (88.1%) |
| Changes in sleep | 662 (8.8%) | 137 (1.8%) | 765 (10.2%) | 628 (82.1%) |
| Hoarseness | 130 (1.7%) | 118 (1.6%) | 200 (2.7%) | 82 (41.0%) |
| Hemoptysis | 133 (1.8%) | 94 (1.3%) | 182 (2.4%) | 88 (48.4%) |
| Night sweats | 480 (6.4%) | 72 (1.0%) | 521 (6.9%) | 449 (86.2%) |
| Lack of appetite | 626 (8.3%) | 59 (0.8%) | 653 (8.7%) | 594 (91.0%) |
| Change in bowel habits | 1465 (19.4%) | 59 (0.8%) | 1491 (19.8%) | 1432 (96.0%) |
| Finger clubbing | 41 (0.5%) | 1 (0.0%) | 41 (0.5%) | 40 (97.6%) |

**Appendix 4. Multivariable analysis of symptoms or signs of cases compared to controls at 1, 3, 6 and 12 months prior to diagnosis/index date**

| Symptom or sign | 12 months OR | 6 months OR | 3 months OR | 1 month OR | At diagnosis OR |
|---|---|---|---|---|---|
| **Finger clubbing** | >1,000 (0.0 - >1,000) | >1,000 (0.0 - >1,000) | >1,000 (0.0 - >1,000) | 60.7 (10.6 - 348.7)*** | 50.1 (8.9 - 283.3)*** |
| **Lymphadenopathy** | 0.7 (0.3 - 1.4) | 1.3 (0.7 - 2.4) | 1.3 (0.8 - 2.3) | 1.7 (1.0 - 2.8)* | 5.8 (3.8 - 8.8)*** |
| **Cough** | 1.9 (1.5 - 2.4)*** | 3.1 (2.4 - 4.0)*** | 4.0 (3.1 - 5.2)*** | 5.0 (3.8 - 6.5)*** | 4.7 (3.5 - 6.3)*** |
| **Hemoptysis** | 2.1 (1.0 - 4.4)* | 3.2 (1.9 - 5.3)*** | 3.1 (1.9 - 4.9)*** | 3.4 (2.2 - 5.4)*** | 3.5 (2.2 - 5.5)*** |
| **Chest crackles or wheeze** | 2.5 (1.9 - 3.5)*** | 3.1 (2.3 - 4.1)*** | 3.0 (2.3 - 4.0)*** | 3.0 (2.3 - 4.0)*** | 3.2 (2.4 - 4.3)*** |
| **Weight loss** | 1.2 (0.9 - 1.8) | 2.1 (1.5 - 2.8)*** | 2.6 (1.9 - 3.4)*** | 2.8 (2.1 - 3.7)*** | 2.9 (2.2 - 3.9)*** |
| **Back pain** | 2.8 (2.1 - 3.6)*** | 2.5 (1.9 - 3.2)*** | 2.5 (1.9 - 3.2)*** | 2.4 (1.9 - 3.1)*** | 2.4 (1.8 - 3.1)*** |
| **Bone pain** | 2.8 (2.1 - 3.7)*** | 2.7 (2.1 - 3.6)*** | 2.4 (1.8 - 3.2)*** | 2.3 (1.7 - 3.0)*** | 2.3 (1.7 - 3.0)*** |
| **Shortness of breath** | 0.7 (0.5 - 1.0)* | 1.0 (0.7 - 1.3) | 1.3 (1.0 - 1.7) | 1.6 (1.2 - 2.1)** | 1.9 (1.4 - 2.5)*** |
| **Fatigue** | 1.6 (1.2 - 2.1)*** | 1.6 (1.3 - 2.1)*** | 1.9 (1.4 - 2.5)*** | 1.8 (1.4 - 2.4)*** | 1.8 (1.3 - 2.3)*** |
| **Chest Pain** | 1.1 (0.8 - 1.4) | 1.2 (0.9 - 1.5) | 1.2 (1.0 - 1.6) | 1.3 (1.0 - 1.6) | 1.4 (1.1 - 1.8)* |
| **Shoulder pain** | 1.3 (0.9 - 1.7) | 1.4 (1.0 - 1.8)* | 1.3 (1.0 - 1.7) | 1.3 (1.0 - 1.7) | 1.3 (0.9 - 1.7) |
| **Ankle swelling** | 1.5 (1.1 - 1.9)** | 1.3 (1.0 - 1.7) | 1.3 (1.0 - 1.7) | 1.3 (1.0 - 1.7) | 1.1 (0.9 - 1.5) |
| **Headache** | 1.0 (0.7 - 1.3) | 1.1 (0.8 - 1.4) | 1.0 (0.8 - 1.3) | 1.0 (0.8 - 1.3) | 1.1 (0.8 - 1.4) |
| **Hoarseness** | 0.9 (0.5 - 1.7) | 1.1 (0.7 - 1.8) | 1.0 (0.6 - 1.6) | 1.1 (0.7 - 1.7) | 1.0 (0.7 - 1.7) |
| **Changes in bowel habits** | 1.2 (0.9 - 1.6) | 1.0 (0.8 - 1.4) | 1.1 (0.8 - 1.5) | 1.0 (0.8 - 1.4) | 1.0 (0.8 - 1.4) |
| **Muscle weakness** | 1.0 (0.7 - 1.3) | 0.9 (0.7 - 1.2) | 1.0 (0.7 - 1.3) | 1.0 (0.8 - 1.3) | 1.0 (0.7 - 1.3) |
| **Night sweats** | 0.9 (0.6 - 1.4) | 0.9 (0.7 - 1.4) | 0.9 (0.7 - 1.3) | 0.9 (0.6 - 1.3) | 0.8 (0.6 - 1.2) |
| **Lack of appetite** | 0.5 (0.3 - 0.7)*** | 0.6 (0.4 - 0.8)** | 0.6 (0.4 - 0.8)** | 0.6 (0.4 - 0.9)** | 0.7 (0.5 - 0.9)* |
| **Dizziness** | 0.8 (0.6 - 1.0) | 0.7 (0.5 - 0.9)** | 0.7 (0.5 - 0.9)** | 0.6 (0.5 - 0.8)** | 0.6 (0.4 - 0.8)*** |
| **Changes in sleep** | 0.8 (0.5 - 1.1) | 0.5 (0.4 - 0.7)*** | 0.4 (0.3 - 0.6)*** | 0.4 (0.3 - 0.6)*** | 0.4 (0.3 - 0.6)*** |
| **Fever** | 0.6 (0.4 - 0.8)*** | 0.5 (0.4 - 0.7)*** | 0.5 (0.4 - 0.6)*** | 0.5 (0.3 - 0.6)*** | 0.4 (0.3 - 0.6)*** |

*Note:* Models adjusted for comorbidities using van Walraven weighted score. Confidence intervals for significant ORs do not incorporate 1.0 due to rounding.

***** p<0.05

****** p<0.01

******* p<0.001

**Appendix 5. Frequency of symptoms and signs in cases and controls with and without chronic respiratory disease**

| Symptom or sign | Chronic respiratory disease | | No chronic respiratory disease | |
|---|---|---|---|---|
| | Control (n=1252) | Case (n=353) | Control (n=5589) | Case (n=345) |
| Cough | 636 (50.8%) | 312 (88.4%) | 1018 (18.2%) | 261 (75.7%) |
| Shortness of breath | 623 (49.8%) | 307 (87.0%) | 990 (17.7%) | 208 (60.3%) |
| Fatigue | 459 (36.7%) | 266 (75.4%) | 1128 (20.2%) | 210 (60.9%) |
| Ankle swelling | 516 (41.2%) | 250 (70.8%) | 1322 (23.7%) | 197 (57.1%) |
| Chest Pain | 439 (35.1%) | 228 (64.6%) | 962 (17.2%) | 175 (50.7%) |
| Chest crackles or wheeze | 307 (24.5%) | 268 (75.9%) | 268 (4.8%) | 129 (37.4%) |
| Back pain | 278 (22.2%) | 191 (54.1%) | 668 (12.0%) | 159 (46.1%) |
| Changes in bowel habits | 337 (26.9%) | 195 (55.2%) | 818 (14.6%) | 141 (40.9%) |
| Muscle weakness | 327 (26.1%) | 177 (50.1%) | 775 (13.9%) | 157 (45.5%) |
| Fever | 433 (34.6%) | 177 (50.1%) | 901 (16.1%) | 145 (42.0%) |
| Weight loss | 165 (13.2%) | 191 (54.1%) | 357 (6.4%) | 117 (33.9%) |
| Headache | 324 (25.9%) | 175 (49.6%) | 881 (15.8%) | 129 (37.4%) |
| Dizziness | 366 (29.2%) | 174 (49.3%) | 953 (17.1%) | 125 (36.2%) |
| Bone pain | 207 (16.5%) | 141 (39.9%) | 518 (9.3%) | 129 (37.4%) |
| Lack of appetite | 142 (11.3%) | 116 (32.9%) | 315 (5.6%) | 80 (23.2%) |
| Shoulder pain | 200 (16.0%) | 92 (26.1%) | 513 (9.2%) | 88 (25.5%) |
| Lymphadenopathy | 35 (2.8%) | 79 (22.4%) | 70 (1.3%) | 72 (20.9%) |
| Night sweats | 113 (9.0%) | 89 (25.2%) | 258 (4.6%) | 61 (17.7%) |
| Changes in sleep | 178 (14.2%) | 90 (25.5%) | 453 (8.1%) | 44 (12.8%) |
| Hemoptysis | 31 (2.5%) | 72 (20.4%) | 36 (0.6%) | 43 (12.5%) |
| Hoarseness | 55 (4.4%) | 45 (12.7%) | 78 (1.4%) | 22 (6.4%) |
| Finger clubbing | 1 (0.1%) | 28 (7.9%) | 1 (0.0%) | 11 (3.2%) |

### Appendix 6. Multivariate analysis of symptoms and signs in patients with and without chronic respiratory disease

| Symptom or sign | Chronic respiratory disease | | | No chronic respiratory disease | | |
|---|---|---|---|---|---|---|
| | Univariate Odds ratio (95%CI) | Multivariate Odds ratio (95%CI) | Multivariate P value | Univariate Odds ratio (95%CI) | Multivariate Odds ratio (95%CI) | Multivariate P value |
| Finger clubbing | 47.3 (6.1 - 364.5) | 17.8 (1.3 - 247.1) | 0.0322 | >1,000 (0.0 - >1,000) | 267.7 (0.1 - >1,000) | 0.1783 |
| Chest crackles or wheeze | 9.4 (6.3 - 14.2)* | 4.9 (2.6 - 9.0) | <0.0001 | 9.8 (7.0 - 13.9)* | 3.2 (2.0 - 5.2) | <0.0001 |
| Hemoptysis | 12.5 (6.2 - 25.3)* | 4.4 (1.7 - 11.5) | 0.0028 | 20.3 (10.2 - 40.5)* | 3.8 (1.5 - 9.8) | 0.0049 |
| Weight loss | 7.1 (4.7 - 10.5)* | 4.0 (2.2 - 7.4) | <0.0001 | 3.8 (2.8 - 5.3)* | 1.6 (1.0 - 2.5) | 0.0643 |
| Lympadenopathy | 7.1 (3.9 - 13.0)* | 3.3 (1.3 - 7.9) | 0.0089 | 12.0 (7.2 - 19.9)* | 8.5 (4.3 - 17.0) | <0.0001 |
| Fatigue | 5.2 (3.6 - 7.6)* | 2.9 (1.6 - 5.5) | 0.0008 | 4.2 (3.2 - 5.6)* | 1.7 (1.1 - 2.6) | 0.0128 |
| Back pain | 4.6 (3.2 - 6.6)* | 2.4 (1.4 - 4.1) | 0.0014 | 4.8 (3.6 - 6.4)* | 2.1 (1.4 - 3.2) | 0.0003 |
| Cough | 6.5 (4.2 - 10.2)* | 2.2 (1.1 - 4.3) | 0.0189 | 12.2 (9.0 - 16.6)* | 6.3 (4.2 - 9.3) | <0.0001 |
| Bone pain | 3.8 (2.6 - 5.5)* | 2.1 (1.1 - 4.0) | 0.0168 | 5.3 (3.9 - 7.2)* | 2.5 (1.6 - 3.9) | 0.0001 |
| Shortness of breath | 6.5 (4.1 - 10.3)* | 1.6 (0.8 - 3.2) | 0.1688 | 5.1 (3.9 - 6.7)* | 1.9 (1.3 - 2.9) | 0.0024 |
| Changes in bowel habits | 2.7 (2.0 - 3.8)* | 1.3 (0.7 - 2.3) | 0.4474 | 2.5 (1.9 - 3.4)* | 0.9 (0.6 - 1.4) | 0.7286 |
| Night sweats | 3.1 (2.1 - 4.7)* | 1.2 (0.6 - 2.4) | 0.5393 | 3.8 (2.6 - 5.7)* | 0.9 (0.5 - 1.7) | 0.8542 |
| Ankle swelling | 2.8 (2.0 - 3.9)* | 1.1 (0.6 - 2.0) | 0.6696 | 3.1 (2.4 - 4.0)* | 1.2 (0.8 - 1.8) | 0.3121 |
| Shoulder pain | 1.6 (1.1 - 2.4) | 1.1 (0.6 - 2.0) | 0.7589 | 2.9 (2.1 - 4.0)* | 1.6 (1.0 - 2.5) | 0.0484 |
| Hoarseness | 2.5 (1.4 - 4.4) | 1.0 (0.5 - 2.3) | 0.9617 | 4.1 (2.2 - 7.7)* | 0.9 (0.4 - 2.2) | 0.8729 |
| Headache | 2.5 (1.9 - 3.5)* | 0.9 (0.5 - 1.7) | 0.8551 | 2.2 (1.7 - 2.9)* | 1.0 (0.7 - 1.6) | 0.8319 |
| Chest Pain | 2.6 (1.9 - 3.6)* | 0.9 (0.5 - 1.6) | 0.7953 | 3.7 (2.8 - 4.8)* | 1.5 (1.0 - 2.2) | 0.0494 |
| Muscle weakness | 2.3 (1.7 - 3.2)* | 0.9 (0.5 - 1.7) | 0.7901 | 3.1 (2.3 - 4.1)* | 1.1 (0.7 - 1.7) | 0.6809 |
| Dizziness | 2.3 (1.7 - 3.3)* | 0.9 (0.5 - 1.6) | 0.7450 | 1.8 (1.3 - 2.4)* | 0.5 (0.3 - 0.8) | 0.0027 |
| Lack of appetite | 2.6 (1.8 - 3.8)* | 0.5 (0.3 - 1.0) | 0.0667 | 1.8 (1.3 - 2.6) | 0.5 (0.3 - 0.9) | 0.0122 |
| Changes in sleep | 1.6 (1.1 - 2.3) | 0.5 (0.3 - 0.9) | 0.0233 | 1.1 (0.7 - 1.6) | 0.3 (0.2 - 0.6) | 0.0004 |
| Fever | 1.6 (1.2 - 2.2) | 0.3 (0.2 - 0.6) | 0.0003 | 2.5 (1.9 - 3.3)* | 0.6 (0.4 - 0.9) | 0.0229 |

*Note:* Models adjusted for comorbidities using van Walraven weighted score

*Significant at p<0.0001
